# Coronavirus and incomes: the COVID-19 pandemic dynamics in Africa in February 2022

**DOI:** 10.1101/2022.04.22.22274058

**Authors:** Igor Nesteruk, Oleksii Rodionov

## Abstract

The relative accumulated and daily characteristics of the COVID-19 pandemic dynamics in Africa were used to find links with the gross domestic product per capita (GDP), percentages of fully vaccinated people and daily numbers of tests per case. A simple statistical analysis of datasets corresponding to February 1, 2022 showed that accumulated and daily numbers of cases per capita, daily numbers of deaths per capita and vaccination levels increase with the increase of GDP. As in the case of Europe, the smoothed daily numbers of new cases per capita in Africa increase with the increasing of the vaccination level. But the increase of the accumulated numbers of cases and daily number of deaths with increasing the vaccination level was revealed in Africa. In comparison with Europe, no significant correlation was revealed between the vaccination level and the number of deaths per case. As in the case of Europe, African countries demonstrate no statistically significant links between the pandemic dynamics characteristics and the daily number of tests per case. It looks that countries with very small GDP are less affected by the COVID-19 pandemic. The cause of this phenomenon requires further research, but it is possible that low incomes limit the mobility of the population and reduce the number of contacts with infected people. In order to overcome the pandemic, quarantine measures and social distance should not be neglected (this also applies to countries with a high level of income and vaccination).

## Introduction

Features of the COVID-19 pandemic dynamics need further study despite the huge number of investigations in this area. In particular, there are many studies comparing the COVID-19 pandemic dynamics in different regions and the impact of various factors [1-18]. Since the populations of different countries are very different, we will use relative characteristics: accumulated numbers of laboratory confirmed cases per capita (CC), averaged daily numbers of new cases per capita (DCC), averaged daily numbers of new deaths per capita (DDC). Since these characteristics change over time, it is worth comparing these relative characteristics at different moments and periods of time [9, 10, 14, 15, 17]. In [9] June 27, 2021 and the following demographic factors: size of population, its density per square kilometer, and the level of urbanization were used to find correlations with CC values in Ukrainian regions and European countries. Since we are already in the third year of the pandemic, it is reasonable to compare the DCC and DDC numbers for the same periods in 2020 and 2021 to find some seasonal and global trends. Since the influence of vaccinations in 2020 can be neglected, the comparison of the COVID-19 pandemic dynamics in 2020 and 2021 enables us to reveal some influences of vaccination levels. In [9] the DCC values for Argentina, Brazil, India, South Africa, Ukraine, EU, the UK, USA, and the whole world for the period from March to August 2020 were compared with the corresponding values in 2021. In [14, 17] DCC and DDC values and vaccination levels in the same countries and Australia for the period from September to January have been compared.

Very interesting results were obtained in [10, 15] for the influence of the vaccination level on the DCC values in Europe and some other countries and regions for two different moments of time. As of September 1, 2021 no statistically significant links were revealed, [10]. As of February 1, 2022, the averaged daily numbers of new cases increased with the increasing vaccination level, [15]. This unexpected result is probably due to the lifting of quarantine restrictions for vaccinated persons who may be ill and transmit the infection more intensely as unvaccinated. Another surprise was close to zero DCC values at low levels of vaccination found for data corresponding to February 1, 2022, [15]. These facts prompted us to investigate the COVID-19 pandemic dynamics in Africa, where the levels of CC and DCC are usually much lower than in Europe and Americas. Thus, we will use the linear regression to reveal the influence of the percentage of fully vaccinated people (VC) and the daily numbers of tests per case (DTC). In this paper we will also try to find statistical links between CC, DCC, DDC, DDC/DCC values and the gross domestic product per capita (GDP) based on purchasing power parity (PPP).

### Data, the linear regression and Fisher test

We will use the data sets regarding the gross domestic product per capita (GDP) based on purchasing power parity (PPP) available in [19] and some COVID-19 characteristics reported by John Hopkins University (JHU) as of February 1, 2022, [20]. The figures corresponding to the versions of files available on February 6, 2022 are presented in Table 1. The characteristics VC, DTC and CC are taken without smoothing. Since daily characteristics DCC (new cases per million) and DDC (new deaths per million) are very random and demonstrate some weekly periodicity, we will use smoothed (averaged) values calculated and displayed by JHU with the use of figures registered during the previous 7 days. This smoothing procedure differs from one proposed in [6, 21, 22], where the values registered in the nearest 7 days were used.

**Table 1.**
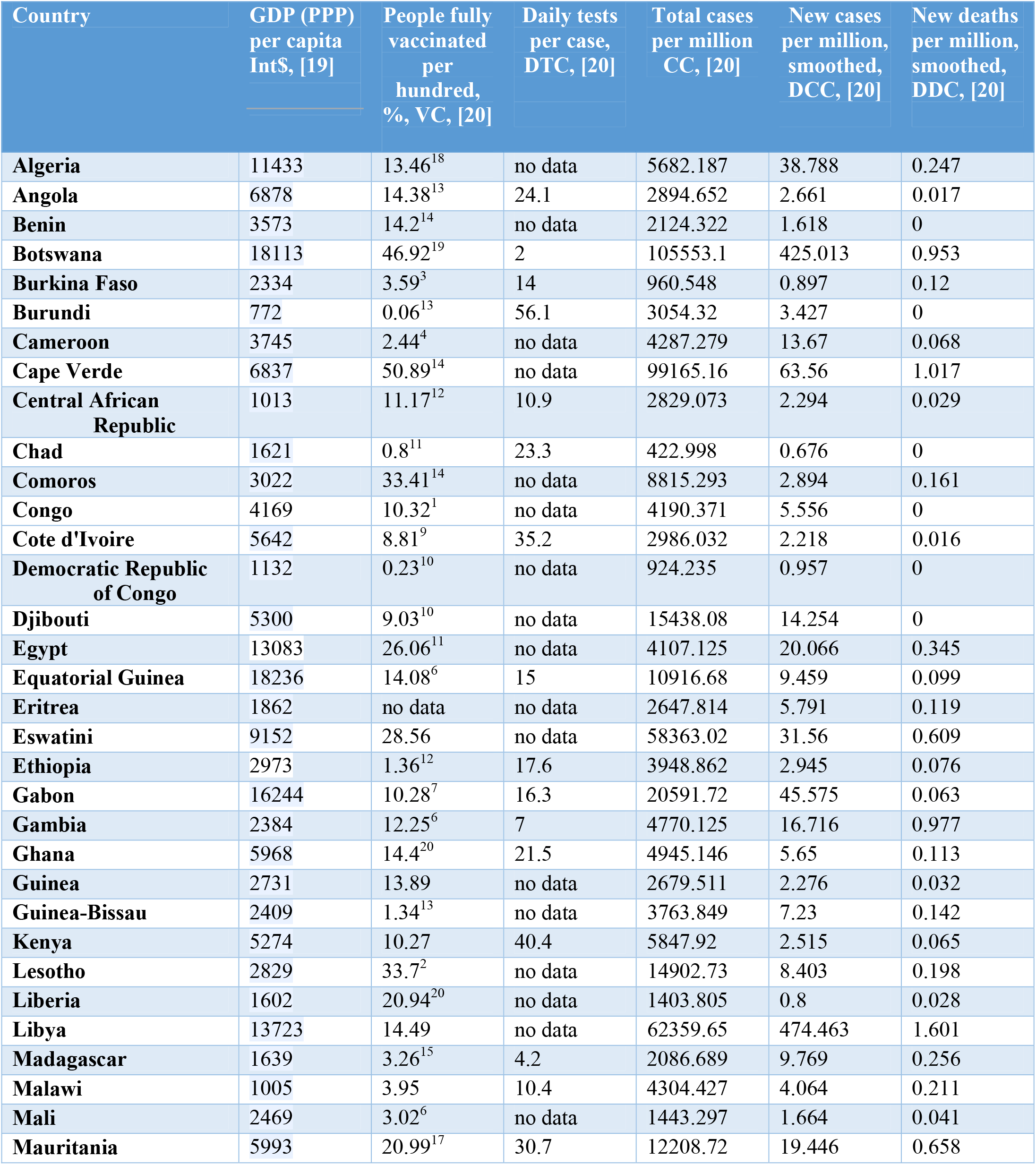
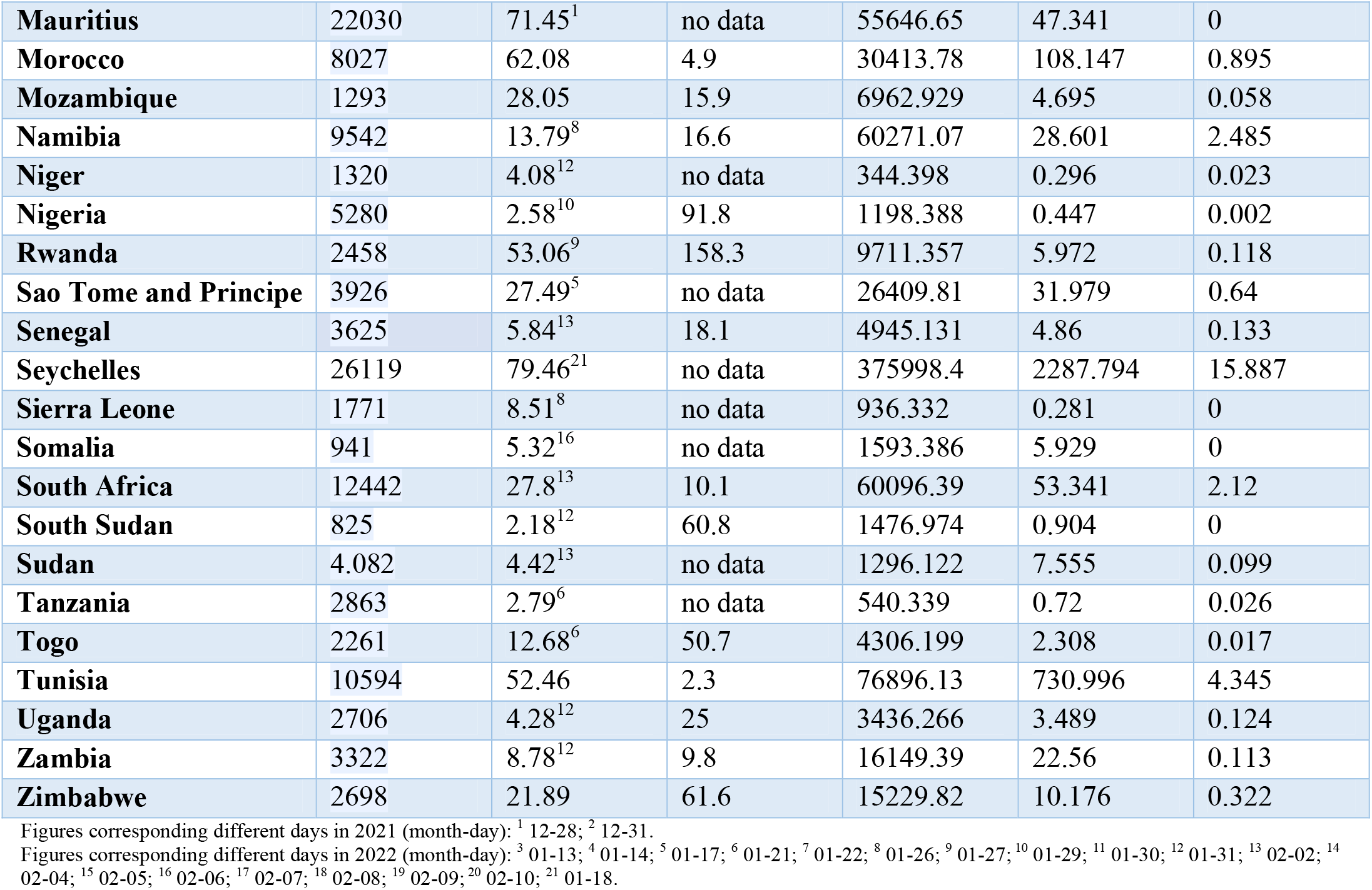
Gross domestic product per capita (GDP) based on purchasing power parity (PPP), accumulated and daily characteristics of the COVID-19 pandemic dynamics in African countries as of February 1, 2022 (figures corresponding to other days are specified in notes).

| Country | GDP (PPP)<br>per capita<br>Int\$, [19] | People fully<br>vaccinated<br>per<br>hundred,<br>%, VC, [20] | Daily tests<br>per case,<br>DTC, [20] | Total cases<br>per million<br>CC, [20] | New cases<br>per million,<br>smoothed,<br>DCC, [20] | New deaths<br>per million,<br>smoothed,<br>DDC, [20] |
| --- | --- | --- | --- | --- | --- | --- |
| Algeria | 11433 | 13.46<sup>18</sup> | no data | 5682.187 | 38.788 | 0.247 |
| Angola | 6878 | 14.38<sup>13</sup> | 24.1 | 2894.652 | 2.661 | 0.017 |
| Benin | 3573 | 14.2<sup>14</sup> | no data | 2124.322 | 1.618 | 0 |
| Botswana | 18113 | 46.92<sup>19</sup> | 2 | 105553.1 | 425.013 | 0.953 |
| Burkina Faso | 2334 | 3.59<sup>3</sup> | 14 | 960.548 | 0.897 | 0.12 |
| Burundi | 772 | 0.06<sup>13</sup> | 56.1 | 3054.32 | 3.427 | 0 |
| Cameroon | 3745 | 2.44<sup>4</sup> | no data | 4287.279 | 13.67 | 0.068 |
| Cape Verde | 6837 | 50.89<sup>14</sup> | no data | 99165.16 | 63.56 | 1.017 |
| Central African<br>Republic | 1013 | 11.17<sup>12</sup> | 10.9 | 2829.073 | 2.294 | 0.029 |
| Chad | 1621 | 0.8<sup>11</sup> | 23.3 | 422.998 | 0.676 | 0 |
| Comoros | 3022 | 33.41<sup>14</sup> | no data | 8815.293 | 2.894 | 0.161 |
| Congo | 4169 | 10.32<sup>1</sup> | no data | 4190.371 | 5.556 | 0 |
| Cote d'Ivoire | 5642 | 8.81<sup>9</sup> | 35.2 | 2986.032 | 2.218 | 0.016 |
| Democratic Republic<br>of Congo | 1132 | 0.23<sup>10</sup> | no data | 924.235 | 0.957 | 0 |
| Djibouti | 5300 | 9.03<sup>10</sup> | no data | 15438.08 | 14.254 | 0 |
| Egypt | 13083 | 26.06<sup>11</sup> | no data | 4107.125 | 20.066 | 0.345 |
| Equatorial Guinea | 18236 | 14.08<sup>6</sup> | 15 | 10916.68 | 9.459 | 0.099 |
| Eritrea | 1862 | no data | no data | 2647.814 | 5.791 | 0.119 |
| Eswatini | 9152 | 28.56 | no data | 58363.02 | 31.56 | 0.609 |
| Ethiopia | 2973 | 1.36<sup>12</sup> | 17.6 | 3948.862 | 2.945 | 0.076 |
| Gabon | 16244 | 10.28<sup>7</sup> | 16.3 | 20591.72 | 45.575 | 0.063 |
| Gambia | 2384 | 12.25<sup>6</sup> | 7 | 4770.125 | 16.716 | 0.977 |
| Ghana | 5968 | 14.4<sup>20</sup> | 21.5 | 4945.146 | 5.65 | 0.113 |
| Guinea | 2731 | 13.89 | no data | 2679.511 | 2.276 | 0.032 |
| Guinea-Bissau | 2409 | 1.34<sup>13</sup> | no data | 3763.849 | 7.23 | 0.142 |
| Kenya | 5274 | 10.27 | 40.4 | 5847.92 | 2.515 | 0.065 |
| Lesotho | 2829 | 33.7<sup>2</sup> | no data | 14902.73 | 8.403 | 0.198 |
| Liberia | 1602 | 20.94<sup>20</sup> | no data | 1403.805 | 0.8 | 0.028 |
| Libya | 13723 | 14.49 | no data | 62359.65 | 474.463 | 1.601 |
| Madagascar | 1639 | 3.26<sup>15</sup> | 4.2 | 2086.689 | 9.769 | 0.256 |
| Malawi | 1005 | 3.95 | 10.4 | 4304.427 | 4.064 | 0.211 |
| Mali | 2469 | 3.02<sup>6</sup> | no data | 1443.297 | 1.664 | 0.041 |
| Mauritania | 5993 | 20.99<sup>17</sup> | 30.7 | 12208.72 | 19.446 | 0.658 |
| **Mauritius** | 22030 | 71.45<sup>1</sup> | no data | 55646.65 | 47.341 | 0 |
| **Morocco** | 8027 | 62.08 | 4.9 | 30413.78 | 108.147 | 0.895 |
| **Mozambique** | 1293 | 28.05 | 15.9 | 6962.929 | 4.695 | 0.058 |
| **Namibia** | 9542 | 13.79<sup>8</sup> | 16.6 | 60271.07 | 28.601 | 2.485 |
| **Niger** | 1320 | 4.08<sup>12</sup> | no data | 344.398 | 0.296 | 0.023 |
| **Nigeria** | 5280 | 2.58<sup>10</sup> | 91.8 | 1198.388 | 0.447 | 0.002 |
| **Rwanda** | 2458 | 53.06<sup>9</sup> | 158.3 | 9711.357 | 5.972 | 0.118 |
| **Sao Tome and Principe** | 3926 | 27.49<sup>5</sup> | no data | 26409.81 | 31.979 | 0.64 |
| **Senegal** | 3625 | 5.84<sup>13</sup> | 18.1 | 4945.131 | 4.86 | 0.133 |
| **Seychelles** | 26119 | 79.46<sup>21</sup> | no data | 375998.4 | 2287.794 | 15.887 |
| **Sierra Leone** | 1771 | 8.51<sup>8</sup> | no data | 936.332 | 0.281 | 0 |
| **Somalia** | 941 | 5.32<sup>16</sup> | no data | 1593.386 | 5.929 | 0 |
| **South Africa** | 12442 | 27.8<sup>13</sup> | 10.1 | 60096.39 | 53.341 | 2.12 |
| **South Sudan** | 825 | 2.18<sup>12</sup> | 60.8 | 1476.974 | 0.904 | 0 |
| **Sudan** | 4.082 | 4.42<sup>13</sup> | no data | 1296.122 | 7.555 | 0.099 |
| **Tanzania** | 2863 | 2.79<sup>6</sup> | no data | 540.339 | 0.72 | 0.026 |
| **Togo** | 2261 | 12.68<sup>6</sup> | 50.7 | 4306.199 | 2.308 | 0.017 |
| **Tunisia** | 10594 | 52.46 | 2.3 | 76896.13 | 730.996 | 4.345 |
| **Uganda** | 2706 | 4.28<sup>12</sup> | 25 | 3436.266 | 3.489 | 0.124 |
| **Zambia** | 3322 | 8.78<sup>12</sup> | 9.8 | 16149.39 | 22.56 | 0.113 |
| **Zimbabwe** | 2698 | 21.89 | 61.6 | 15229.82 | 10.176 | 0.322 |

Figures corresponding different days in 2021 (month-day): <sup>1</sup> 12-28; <sup>2</sup> 12-31.
Figures corresponding different days in 2022 (month-day): <sup>3</sup> 01-13; <sup>4</sup> 01-14; <sup>5</sup> 01-17; <sup>6</sup> 01-21; <sup>7</sup> 01-22; <sup>8</sup> 01-26; <sup>9</sup> 01-27; <sup>10</sup> 01-29; <sup>11</sup> 01-30; <sup>12</sup> 01-31; <sup>13</sup> 02-02; <sup>14</sup> 02-04; <sup>15</sup> 02-05; <sup>16</sup> 02-06; <sup>17</sup> 02-07; <sup>18</sup> 02-08; <sup>19</sup> 02-09; <sup>20</sup> 02-10; <sup>21</sup> 01-18.

We will use the linear regression to calculate the regression coefficients *r* and the coefficients *a* and *b* of corresponding best fitting straight lines, [23]:

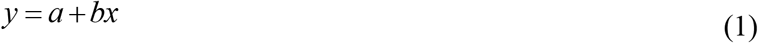

where *x* are GDP, VC, DTC values and *y* are DCC, DDC, DDC/DCC, and CC values.

We will also use the F-test for the null hypothesis that says that the proposed linear relationship (1) fits the data sets. The experimental values of the Fisher function can be calculated with the use of the formula:

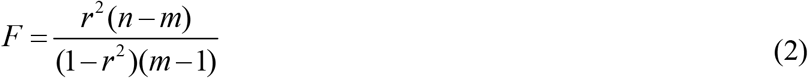

where *n* is the number of observations (number of countries taken for statistical analysis); *m*=2 is the number of parameters in the regression equation, [23]. The corresponding experimental values *F* have to be compared with the critical values *F*_*C*_ (*k*_1_, *k*_2_) of the Fisher function at a desired significance or confidence level (*k*_1_ = *m* −1, *k*_2_ = *n* − *m*, see, e.g., [24]). If *F* / *F*_*C*_ (*k*_1_, *k*_2_) < 1, the null hypothesis is not supported by the results of observations. The highest values of *F* / *F*_*C*_ (*k*_1_, *k*_2_) correspond to the most reliable hypotheses (see, e.g., [25]).

## Results and discussion

The results of the linear regression application are presented in Table 2 and Figs. 1-4. Due to the lack of some data, the numbers of observations *n* (numbers of countries taken for statistical analysis) are different for different relationships. Corresponding values of *n*, the regression coefficients *r*, coefficients *a* and *b*; values of the Fisher function *F, F*_*C*_ (*k*_1_, *k*_2_) and *F* / *F*_*C*_ (*k*_1_, *k*_2_) are shown in Table 2. The best fitting lines (1), calculated with the use of corresponding values *a* and *b* are shown in Figs. 1-4 by blue, black, red, magenta, green and yellow solid lines for the DCC, DDC, DDC/DCC, CC, DTC and VC values, respectively. The same colors we use for values listed in Table 1 and represented by “crosses”.

**Table 2.** Optimal values of parameters in eq. (1), correlation coefficients and the results of Fisher test applications.

| Number of Figure, relationship | Number of observations $n$ | Correlation coefficient $r$ | Optimal values of parameters $a$ in eq. (1) | Optimal values of parameters $b$ in eq. (1) | Experimental value of the Fisher function $F$, eq. (2), $m=2$ | Critical value of Fisher Function $F_c(I, n-2)$ for the confidence level 0.1, [9] | $F/F_c$ |
| --- | --- | --- | --- | --- | --- | --- | --- |
| 1, DCC versus GDP | 54 | 0.6001 | -110.1448 | 0.0341 | 29.2670 | 2.81 | 10.42 |
| 1, DDC versus GDP | 54 | 0.566 | -0.5881 | 2.1806e-004 | 24.5389 | 2.81 | 8.73 |
| 1, DDC/DCC versus GDP | 54 | -0.1945 | 0.0254 | -8.3715e-007 | 2.0454 | 2.81 | 0.73 |
| 1, CC versus GDP | 54 | 0.6869 | -1.4432e+004 | 6.5128 | 46.4608 | 2.81 | 16.5 |
| 1, DTC versus GDP | 29 | -0.2841 | 39.5219 | -0.0018 | 2.3711 | 2.91 | 0.81 |
| 1, VC versus GDP | 53 | 0.6254 | 6.1688 | 0.0020 | 32.7550 | 2.82 | 11.62 |
| 2, 3, DCC versus VC | 53 | 0.5615 | -90.3101 | 9.8639 | 23.4885 | 2.82 | 8.33 |
| 2, 3, DDC versus VC | 53 | 0.5507 | -0.5054 | 0.0655 | 22.2044 | 2.82 | 7.87 |
| 2, 3, DDC/DCC versus VC | 53 | -0.1358 | 0.0239 | -1.8064e-004 | 0.9587 | 2.82 | 0.34 |
| 2, 3, CC versus VC | 53 | 0.6720 | -1.2063e+04 | 1.9665e+003 | 41.9993 | 2.82 | 14.89 |
| 2, 3, DTC versus VC | 29 | 0.0889 | 26.6944 | 0.1705 | 0.2149 | 2.91 | 0.074 |
| 4, DCC versus DTC | 29 | -0.2604 | 88.6188 | -1.2171 | 1.9643 | 2.91 | 0.68 |
| 4, DDC versus DTC | 29 | -0.2908 | 0.7479 | -0.0085 | 2.4946 | 2.91 | 0.86 |
| 4, DDC/DCC Versus DTC | 29 | -0.1730 | 0.0287 | -1.5467e-004 | 0.8330 | 2.91 | 0.29 |
| 4, CC versus DTC | 29 | -0.2895 | 2.3301e+004 | -229.7286 | 2.4693 | 2.91 | 0.85 |

**Fig. 1.**
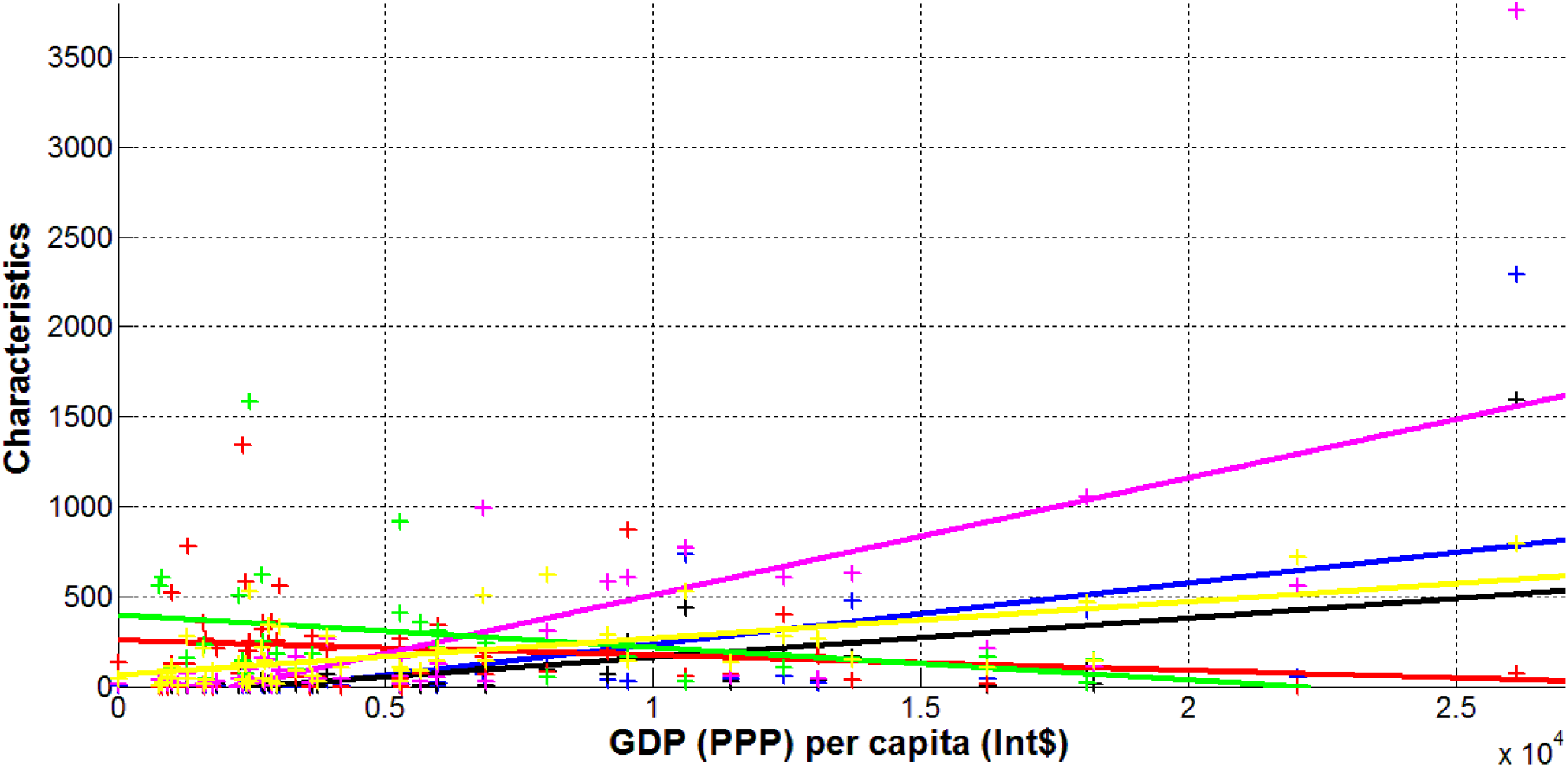
Characteristics of the COVID-19 pandemic in Africa versus gross domestic product based on purchasing power parity GDP (PPP) per capita in international US dollars. New cases per million (DCC, blue), new deaths per 100 million (DDC*100, black), daily mortality rate (DDC*10^4^/DCC, red), accumulated number of cases per 10,000 (CC/100, magenta), daily tests per case ratio (multiplied by 10, DTC*10, green), numbers of fully vaccinated people per thousand (VC*10, yellow). “Crosses” represent the datasets corresponding to February 1, 2022 (Table 1), best fitting lines (1) are solid.

Table 2 and Fig. 1 illustrate that DCC, DDC, CC and VC values increase with the increasing of the GDP. Moreover, the numbers of cases and deaths can tend to zero at GDP < 2,000 USD (see blue, black and magenta lines in Fig. 1). Characteristics DDC/DCC and DTC show no statistically significant correlation with GDP, since *F* / *F*_*C*_ (*k*_1_, *k*_2_) < 1 for these cases (see the last column in Table 2). Thus, richer African countries have more COVID-19 cases and deaths per capita despite the higher level of vaccination. These unexpected results require further research, but it is possible that low incomes limit the mobility of the population and reduce the number of contacts with infected people.

The increase of vaccination level VC increases the accumulated numbers of cases per capita CC and daily characteristics DCC and DDC (see positive values of the parameter *b* in Table 2, magenta, blue and black lines in Figs. 2 and 3). This unexpected fact needs some discussion. First, we should pay attention to the rather high value of the correlation coefficient for the dependence of the vaccination level VC on the per capita income GDP (*r*=0.6869 is the highest value among all dependences versus GDP, see Table 2). On the other hand, for the dependences versus VC, the *F* / *F*_*C*_ (*k*_1_, *k*_2_) values are lower than the corresponding values for the dependences versus GDP (see Table 2). Thus, the increase of the CC, DCC and DDC figures in more vaccinated countries can be explained by the fact that these countries are richer and have a higher mobility of population. We must also take into account that quarantine restrictions were removed for vaccinated persons, who are nevertheless able to be infected and to spread the virus in crowded places.

**Fig. 2.**
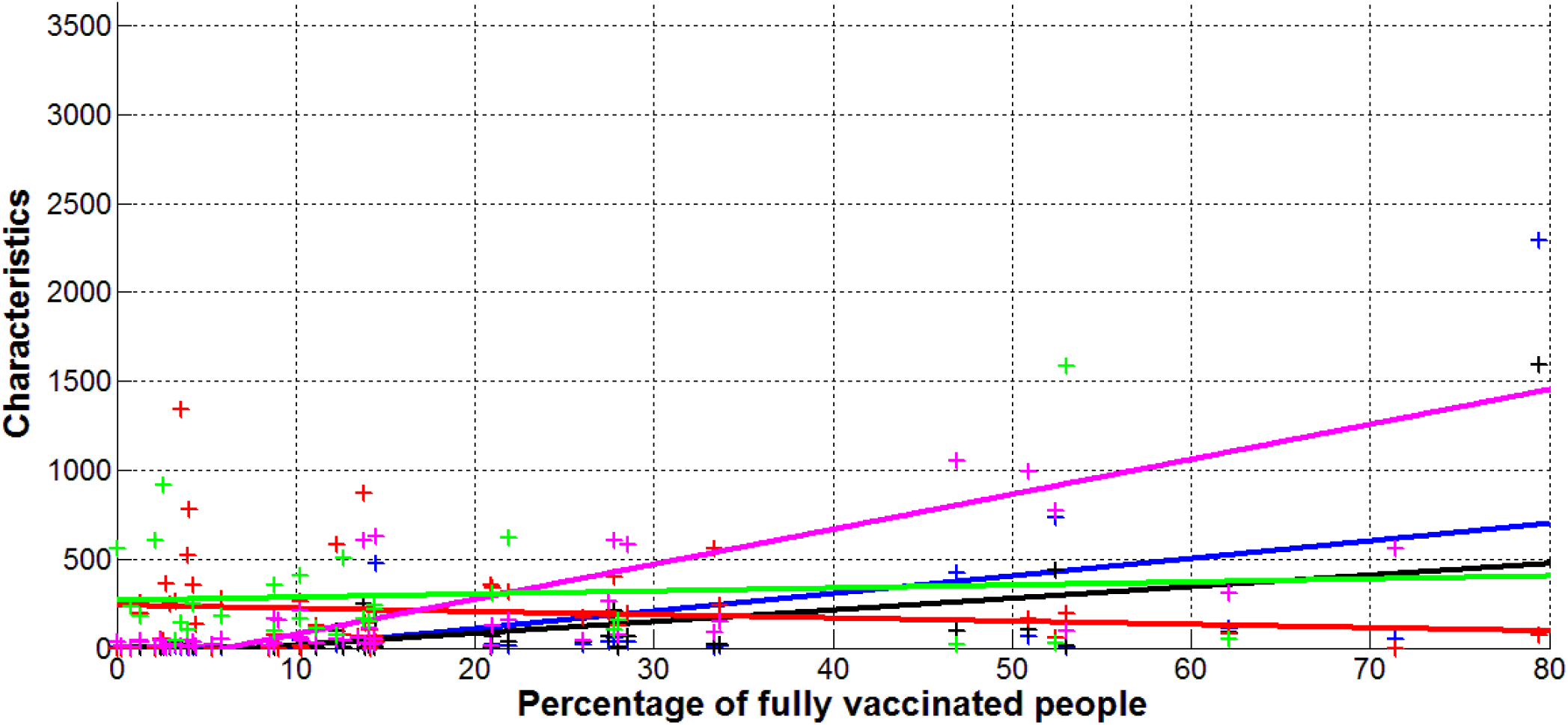
Characteristics of the COVID-19 pandemic in Africa versus percentage of fully vaccinated people. New cases per million (DCC, blue), new deaths per 100 million (DDC*100, black), daily mortality rate (DDC*10^4^/DCC, red), accumulated number of cases per 10,000 (CC/100, magenta), daily tests per case ratio (multiplied by 10, DTC*10, green). “Crosses” represent the datasets corresponding to February 1, 2022 (Table 1), best fitting lines (1) are solid for Africa.

**Fig. 3.**
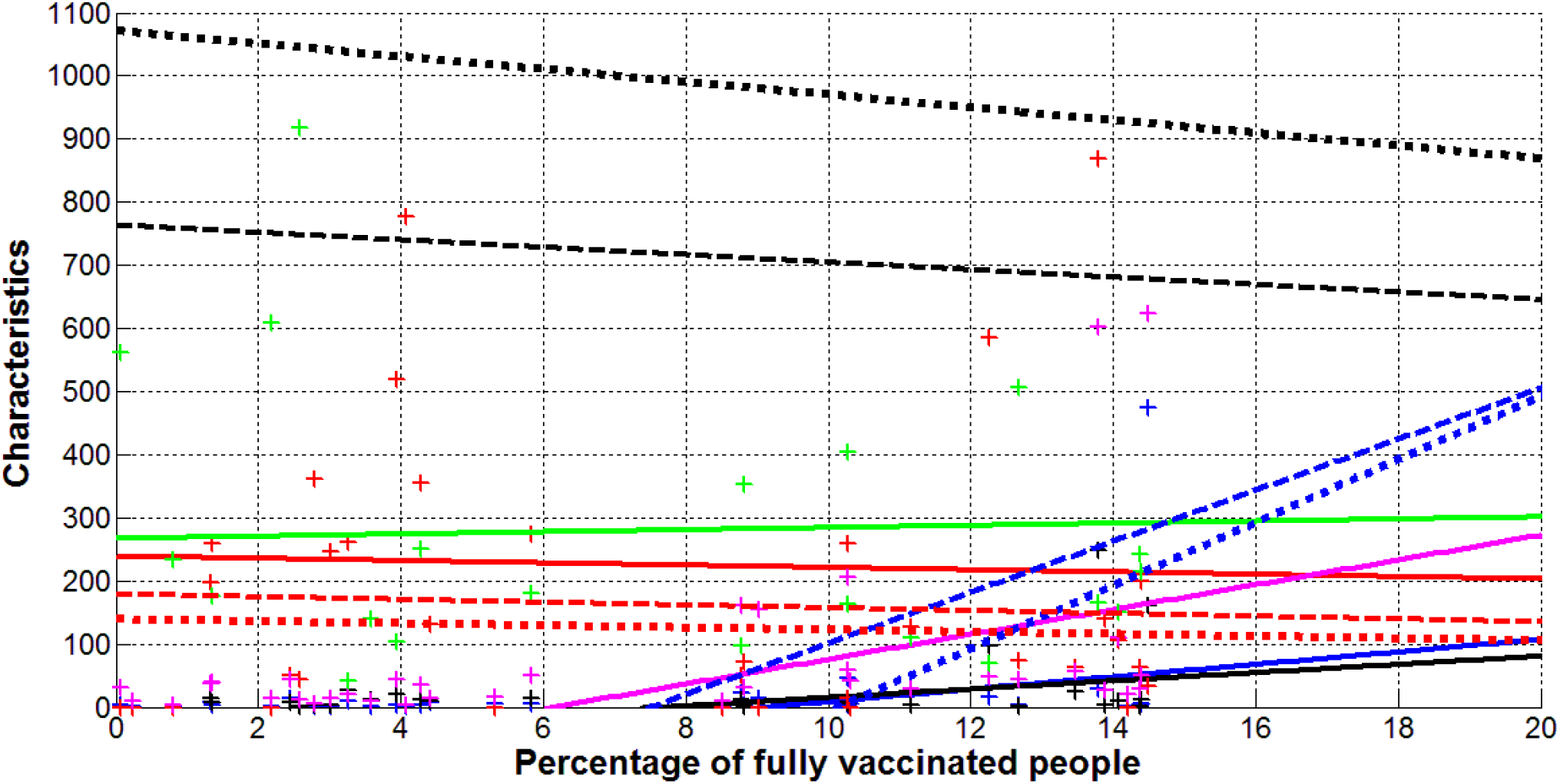
Comparison of the COVID-19 pandemic in Africa, Europe and the world. New cases per million (DCC, blue), new deaths per 100 million (DDC*100, black), daily mortality rate (DDC*10^4^/DCC, red), accumulated number of cases per 10,000 (CC/100, magenta), daily tests per case ratio (multiplied by 10, DTC*10, green). “Crosses” represent the datasets for Africa corresponding to February 1, 2022 (Table 1), best fitting lines (1) are solid for Africa, dotted for Europe [15], dashed for complete data set listed in [15].

In Fig. 3, the best fitting lines for characteristics DCC, DDC, and DDC/DCC in Africa (solid blue, black red lines respectively) are compared with the corresponding lines for Europe (dotted) and the world (dashed). The parameters *a* and *b* for corresponding equations (1) have been calculated in [15] with the use of JHU datasets [20] for the same day (February 1, 2022). The daily numbers of new cases are much lower in Africa than in Europe (compare blue solid and dotted lines in Fig. 3). In both continents and globally, the DCC figures are higher for more vaccinated countries and tend to zero at VC < 8% (see blue lines in Fig. 3). Due to the close correlation between VC and GDP values, we can conclude that probably poor countries have suffered less from the COVID-19 pandemic. As mentioned earlier, this can be explained by lower mobility of poor people, which reduces the number of contacts. We should also not forget the large numbers of undetected cases (see, e.g., [21, 26-31]), which may be higher in poor countries.

In comparison with Europe, the numbers of deaths per capita in Africa is much lower (compare black dotted and solid lines in Fig. 3). We can also see opposite trends for DDC values versus vaccination level. The increase in DDC figures in more vaccinated African countries can be related to higher incomes and mobility. As in Europe, the mortality rate DDC/DCC in Africa decreases with the increase of the vaccination level (see bold red lines in Figs. 2 and 3) but this relationship is not statistically significant for Africa (for Europe, the value *F* / *F*_*C*_ (*k*_1_, *k*_2_) =6.19 has been calculated in [15]). The higher levels of mortality rate DDC/DCC in Africa (compare red lines in Fig. 3) can be probably connected with lower levels of the COVID-19 patient treatment.

Fig. 4 illustrates that CC, DCC, DDC, and DDC/DCC values decrease with the increase of the daily tests per case ratio. The same trend was revealed for the accumulated numbers of cases per capita in Europe, [15]. But no statistically significant correlations were revealed both in Africa (see last four rows of Table 2) and Europe (see [15]). Nevertheless, very high DTC values enable to control the pandemic completely (corresponding cases of Australia and Hong Kong were discussed in [10, 15]).

**Fig. 4.**
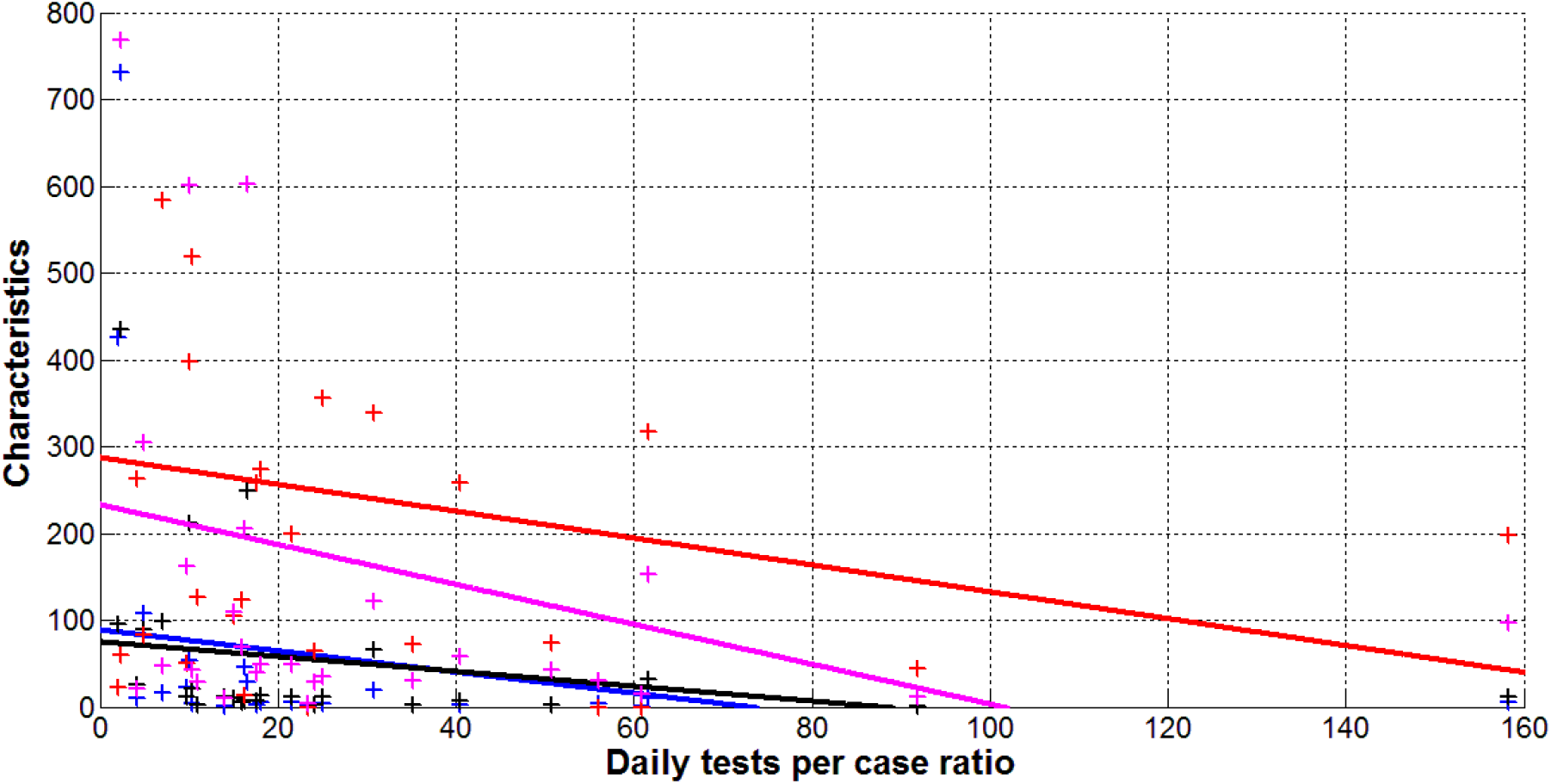
Characteristics of the COVID-19 pandemic in Africa versus daily tests per case ratio. New cases per million (DCC, blue), new deaths per 100 million (DDC*100, black), daily mortality rate (DDC*10^4^/DCC, red), accumulated number of cases per 10,000 (CC/100, magenta) “Crosses” represent the datasets corresponding to February 1, 2022 (Table 1), best fitting lines (1) are solid

## Conclusions

A simple statistical analysis of relative accumulated and daily characteristics of the COVID-19 pandemic in Africa (calculated as of February 1, 2022) revealed that the numbers of cases and deaths per capita increase for richer countries. The high income levels probably increase the mobility and the number of contacts. This leads to an increase in new cases and deaths. Therefore, in order to overcome the pandemic, quarantine measures and social distance should not be neglected (this also applies to countries with a high level of income and vaccination).

## Data Availability

All data produced in the present work are contained in the manuscript

